# Nine-month presence of SARS-CoV-2 in saliva: a case report

**DOI:** 10.1101/2021.10.08.21262462

**Authors:** Gijsbert J. Jansen, Marit Wiersma

## Abstract

SARS-CoV-2 is a novel coronavirus that mainly affects the upper airways. Approximately one third of all detected cases is asymptomatic. We report an asymptomatic individual who tested positive for SARS-CoV-2 over a period of nine months. Of this individual, whole mouth saliva was tested by a novel fluorescence *in situ* hybridization-based assay which detects only the ‘active’ form of the virus. During the observation period of nine months, there was a possible co-infection with a second SARS-CoV-2 variant accompanied by none or very low antibody production until the possible co-infection. We suspect that the SARS-CoV-2 infection in this individual is limited to the salivary glands and does not spread (much) throughout the systemic compartment(s) of the body.

## Introduction

SARS-CoV-2, causing the COVID-19 pandemic, infected over 235.175.000 persons by October 2021 and caused more than 4.800.000 deaths worldwide.^1^ This novel coronavirus mainly affects the upper respiratory system, with the most reported symptoms being flu-like, such as fever, cough and myalgia or fatigue.^2^ However, symptoms are inconsistent and differ greatly between patients. Moreover, approximately one third of SARS-CoV-2-infected persons appear to be asymptomatic.^3,4^ Here, we report a case of an asymptomatic individual who tested positive for SARS-CoV-2 in whole mouth saliva since January 2021.

## Methods

The Fluorescence *In Situ* Hybridization (FISH)-based SARS-CoV-2 test on saliva^5^ was performed as follows: 5 ml of saliva was mixed with 5 ml Biotrack Preservation Fluid and fixed for 10 minutes at room temperature. The fixed cell suspension was mixed 1:2 (50 µl : 100 µl) with the Biotrack COV19 Probe. This mixture was hybridized at 50°C for 90 minutes and, thereafter, washed with 1 ml Biotrack Wash Buffer for 30 minutes at 60°C. The mixture was centrifuged (10 minutes at 800 x g), the supernatant was removed, the pellet resuspended with 50 µl Milli-Q and 10 µl was spotted onto a microscopic slide. A Leica DM2500 fluorescence microscope equipped with a Leica EL6000 mercury lamp, a Leica DFC450C camera and the software Leica application suite V4.20 was utilized to capture images.

The Wantai SARS-CoV-2 Ab ELISA, according to the manufacturer’s instructions, was utilized to test for SARS-CoV-2 antibody presence.

### Case

A healthy male in his 50’s was presented to our research laboratory for our Fluorescence *In Situ* Hybridization (FISH)-based SARS-CoV-2 test on saliva^5^ in December 2020. This was preliminary meant to be a screening test, as one of his colleagues was tested positive for SARS-CoV-2 by RT-PCR and the subject displayed no symptoms. He tested negative for SARS-CoV-2 using the FISH-based molecular test.^5^ However, in January 2021, the man lost both taste and smell and returned to our laboratory. Again, he tested negative for SARS-CoV-2. Due to the specificity of his complaints, there was a strong presumption for SARS-CoV-2. The saliva sample was concentrated 20x, which lead to a very slight positive result for SARS-CoV-2 particles. The next day, he tested very strongly positive for SARS-CoV-2 in saliva. Up till the end of September 2021, the subject kept testing positive for SARS-COV-2 in his saliva, of which 95% of the time strongly positive (Figure 1) and 5% of the time slightly positive.

**Figure 1:**
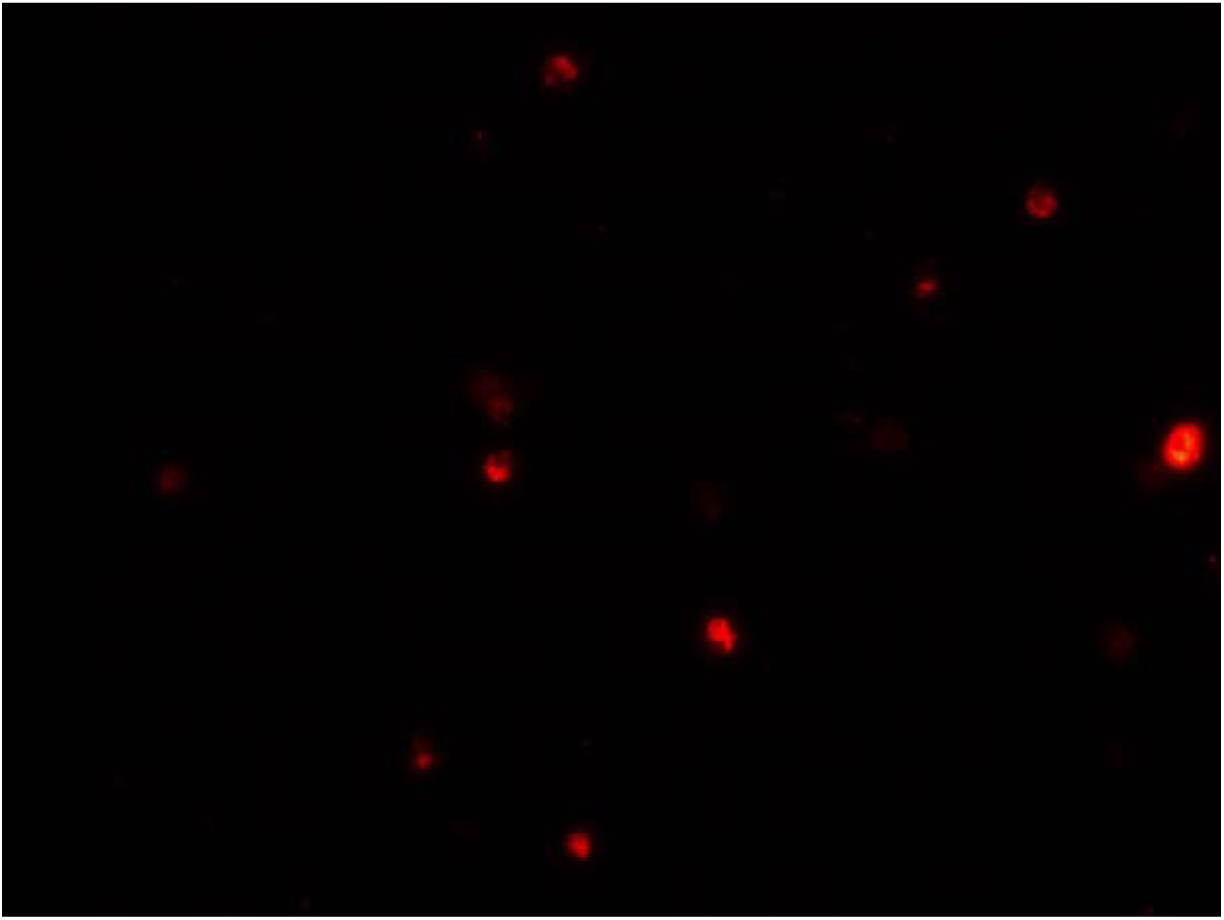
A strong positive image. The saliva sample is FISHed with a SARS-CoV-2 specific probe with a Cy3 label^5^. Infected white blood cells in the sample show different amounts of virus particles present, which correlates with the intensity of the Cy3 in the image.

A RT-PCR test of a nasopharyngeal swab was performed twice during this time period, but was always negative for SARS-CoV-2. Besides the loss of smell and taste, no other symptoms were present until the beginning of July 2021. At that time, the subject presented the following symptoms: fever, cough, nasal congestion, headache, muscle pain, fatigue and loss of smell and taste. We suspect that the subject was (co-)infected with the delta-variant through a relative (confirmed infection by RT-PCR), as that variant had the highest prevalence in the Netherlands at that time.

From March 2021, the presence of antibodies against SARS-CoV-2 was tested once a month utilizing the Wantai SARS-CoV-2 Ab ELISA test. In March, April and May, no antibodies were detected. In June, antibodies were present, but were just above the detection limit. However, in July, before the beginning of the above-mentioned symptoms, no antibodies were detected anymore. In August and September, we could detect antibodies well above the detection limit. The subject was not vaccinated during this time period.

## Discussion

The case presented here is interesting as it describes a subject that tested positive for SARS-CoV-2 in whole mouth saliva by a FISH-based method for 9 months. The RT-PCR on nasopharyngeal swab, performed twice during this period, was found to be negative for SARS-CoV-2. Moreover, none or a very low level of antibodies against SARS-CoV-2 were detected before August 2021. Antibodies were detected in August and September, after a possible (co-)infection with multiple symptoms present.

The test utilized in this case report is a FISH-based test.^5^ FISH is a rapid, reliable and easy method with a high sensitivity for detection of specific RNA of DNA sequences that has been used for more than 20 years.^6,7^ The FISH-probe utilized is SARS-CoV-2-specific and fluorescently-labeled with Cy3. This probe is designed specifically to detect the intracellular antisense genomic RNA of the nucleocapsid gene of SARS-CoV-2, which is the replicative form of SARS-CoV-2.^5^ Therefore, this FISH-based method only detects ‘active’ virus particles and not remnants of an earlier infection.

The choice for the utilization of whole mouth saliva as a matrix instead of a nasopharyngeal swab is due to the following reasons: saliva collection is a less invasive technique, is easy to collect and no (trained) personnel is necessary, which reduces the infection risk.^8^ As multiple viruses, including Epstein-Barr Virus, norovirus and influenza A/B virus, can be detected in saliva^9^, it is no surprise that this is also the case for SARS-CoV-2. In the last few months, the use of saliva as a test matrix for SARS-CoV-2 infection has gotten increased attention. More so, as SARS-CoV-2 was shown to be present in the cells of the salivary glands.^8,10^ Interesting for this case study was a study that showed that SARS-CoV-2 particles may only be present in the salivary glands in the early stage of the infection.^8,10,11^ From the salivary glands, the virus may then spread throughout the body. This may result in a positive SARS-CoV-2 saliva test, but a negative RT-PCR, as the virus is not yet present in the nose and/or throat.^10,11^ An infection with SARS-CoV-2 of the salivary glands may also explain the loss of taste and smell in patients. Another study suggested that asymptomatic patients have SARS-CoV-2 mostly in their salivary glands and that infected saliva is the transmission route from these patients.^12^ These studies confirm our observations in this case report: an asymptomatic patient where the presence of SARS-CoV-2 can only be found in saliva.

Next to SARS-CoV-2 mostly being present in the salivary glands of asymptomatic individuals, it was shown that asymptomatic individuals had decreased SARS-CoV-2 IgG levels in the acute phase of the disease when compared to symptomatic individuals. Moreover, asymptomatic individuals have a higher chance on becoming seronegative for SARS-CoV-2 IgG antibodies after an infection compared to symptomatic individuals.^13^ This explains our finding of none or very low presence of antibodies until August in this case report.

One striking observation at the end of the 9-month period was the upsurge of symptoms. We suspect that this was caused by a co-infection of another SARS-CoV-2 variant, probably the delta variant as this variant was at that time the most present in the Netherlands, but this assumption was not confirmed by accompanying laboratory work. There are several news bulletins and a couple scientific articles in which a co-infection with 2 variants of SARS-CoV-2 is described.^14,15^ Thus, it seems possible that the subject described in this case report indeed suffered a co-infection of 2 variants of SARS-CoV-2.

In conclusion, our presentation of this case of SARS-CoV-2 infection has suggested that the virus may reside in the salivary glands for a long time without giving rise to any symptoms, but also not developing immunity. We suspect that the SARS-CoV-2 infection in this individual is limited to the salivary glands and does not spread (much) throughout the body. Further research is needed to establish whether this is a single case or more individuals have this type of infection. Also, the contagiousness of individuals which harbor SARS-CoV-2 particles in their saliva glands during extended periods of time is unknown and may be indicative for a yet unidentified reservoir of SARS-CoV-2 spreading individuals.

## Data Availability

Data are available upon request

## Acknowledgements

The authors wish to thank Annika Gorter and Lucinda Asraf for assistance with the analyses.

## Author contribution

GJJ and MW collected patient data, carried out analyses and drafted, reviewed and revised the manuscript.

## Ethical approval

This study was approved by the Medical ethics Committee of Isala Clinics, Zwolle, The Netherlands (METc 211004).

## Informed consent statement

Informed consent was obtained from the subject involved in the study.

